# Integrating Multilevel, Multidomain and Multimodal Neuroimaging Factors to Predict Early Alcohol Exposure Trajectories Using Explainable AI

**DOI:** 10.1101/2025.03.12.25323356

**Authors:** Ana Ferariu, Hansoo Chang, Ashni Kumar, Alexandra Sahl, Stephanie Gorka, Lei Wang, Wesley K. Thompson, Fengqing Zhang

## Abstract

Alcohol consumption tends to increase from childhood to adolescence, and risk factors at the individual, family, and environmental level (multilevel, multidomain factors), as well as changes in brain structure and function, have been associated with the likelihood of developing alcohol use disorder (AUD) or binge drinking later in life. Most studies have focused on limited subsets of multilevel or neuroimaging factors, typically emphasizing the risk of alcohol initiation, binge drinking, or AUD in cross-sectional designs rather than exploring longitudinal alcohol consumption trajectories. Our study addresses these gaps by examining a comprehensive set of multilevel, multidomain factors and multimodal brain imaging features to prospectively predict early alcohol sipping trajectories over time with large data from the Adolescent Brain Cognitive Development Study. We applied machine learning methods to baseline individual, family, and environmental factors as well as structural and functional brain connectivity features, analyzing these predictors separately and in combination.

Key findings reveal that functional connectivity and multilevel factors distinguish youth with an increasing alcohol sipping trajectory from those who initially experimented with alcohol but reduced their consumption over time. Moreover, we found important structural and functional features that predicted those who increasingly sipped over time versus the ones who did not engage in alcohol experimentation. Stable interactions between age, socioeconomical status and positive attitudes towards drinking could predict a pattern of increasing alcohol sipping over time. These trends could inform how individual, family, and environmental factors together with brain imaging features impact the development of different alcohol sipping trajectories over time.

## 1. Introduction

Alcohol use is widely prevalent and has been shown to be associated with negative outcomes at the individual, familial and societal level (Bryant et al., 2003; Lees et al., 2020; Sudhinaraset et al., 2016; Watts et al., 2023). Exposure to alcohol during neurodevelopment can disrupt neural pathways and impair cognitive abilities, potentially increasing the likelihood of engaging in problematic drinking or developing alcohol use disorder (AUD) (Lees et al., 2020; May et al., 2022). However, it remains unclear how the interplay of different factors, such as demographics, genetics, culture, environment, physical health and mental health, impacts vulnerability to early alcohol exposure and ultimately, the risk of problematic drinking and alcohol misuse.

The Social-Ecological Model provides a useful framework for examining these influences at multiple levels, individual, interpersonal, organizational, community, and societal, highlighting how personal and environmental factors collectively shape alcohol-related behaviors (Gruenewald et al., 2014). Individual-level factors, such as being male (Squeglia & Cservenka, 2017) or White (Sudhinaraset et al., 2016), having a sedentary lifestyle (Korhonen et al., 2009; Nagata et al., 2024; Purba et al., 2023) and disturbed circadian rhythm (Hasler et al., 2015), as well as increased impulsivity (Aloi et al., 2020; López-Caneda et al., 2013; Stautz & Cooper, 2013) and psychopathological problems (Meque et al., 2019), were associated with higher risk of alcohol exposure and increased risk of alcohol use problems later in life. At the family and community levels, socio-economic status (SES) can play a significant role in shaping alcohol consumption patterns (Huckle et al., 2010; Wahlström et al., 2023; Wells & Östberg, 2018). Close family attachment, negative attitudes towards drinking, and the establishment of rules and penalties regarding alcohol use within the household, as well as less life stress associated with residential stability, have been linked to reduced alcohol consumption (Hahm et al., 2003; Hallihan et al., 2023). Genetic factors and prenatal substance use have adverse health outcomes and can negatively impact brain structure and function, and subsequently, alcohol use behaviors (Gu et al., 2024). While most studies have focused on a small subset of these factors, it remains unclear how these multilevel multidomain factors collectively and interactively impact the patterns of early alcohol exposure over time as individuals transition from childhood to early adolescence. Studies focusing on brain biology using neuroimaging have shown alterations of brain structure and function in different levels of alcohol exposure. Several studies have investigated neuroanatomical predictors of alcohol consumption in adolescents, revealing some inconsistencies in their findings (Honarvar et al., 2023). However, gray matter and white matter volume, surface area and cortical thickness in multiple cortical and subcortical regions have been shown to play a role in the prediction of future problematic drinking episodes in adolescents (Honarvar et al., 2023). At the functional level, alterations in many functional networks, as well as between functional networks have been linked to alcohol misuse in adolescents and adults (Morales et al., 2021; Suk et al., 2021). Resting state hyperconnectivity in networks associated with emotion, reward, executive control and default mode was associated with early binge drinking behavior and alcohol use severity (DeWitt et al., 2014; Fede et al., 2019; Morales et al., 2021; Norman et al., 2011). Resting-state functional connectivity (rs-FC) prior alcohol initiation may be an impactful factor in risky alcohol consumption. Despite evidence of changes in brain structure and function in alcohol-using adolescents, there is a greater need for information about abnormalities in brain structure and functional connectivity prior to alcohol initiation and to problematic alcohol use behaviors in longitudinal frameworks (Feldstein Ewing et al., 2014).

Multilevel, multidomain features and brain biology do not operate in isolation, but the extent to which brain structure and function, together with multilevel, multidomain factors, jointly and interactively impact alcohol intake remains unclear. Findings from hypothesis-driven studies have shown that genetic factors, i.e. family history of AUD (Cservenka et al., 2014; Gu et al., 2024), psychopathology, i.e. childhood trauma (Silveira et al., 2020) and lifestyle factors, i.e. cumulative life stress (Casement et al., 2014) could impact executive dysfunction which could possibly predict future high-risk drinking in adolescents. These findings highlight that risk for early alcohol exposure and problematic drinking is not driven by a single factor, but by a synergistic interplay of biological, psychological and social factors. Identifying these nuanced relationships early is important for developing targeted intervention and prevention strategies. While previous studies have typically focused on older adolescents, were conducted cross-sectionally and captured only a snapshot of alcohol use, i.e. alcohol initiation, binge drinking or AUD, particularly within older cohorts, there remains a critical need for more comprehensive insights into how these multilevel, multidomain factors, along with multimodal brain imaging features, shape longitudinal alcohol consumption patterns during neurodevelopment (Hahm et al., 2003; Hallihan et al., 2023; Huckle et al., 2010; Squeglia & Cservenka, 2017; Wells & Östberg, 2018).

Our current study aimed to bridge the limitations found in the literature by addressing a younger baseline age (mean age = 9.92 years), adopting a longitudinal approach from childhood to early adolescence, and investigating longitudinal alcohol sipping behavior as a specific outcome. To our knowledge, this was one of the first studies to explore multi-modal brain imaging in 9-10-year-old adolescents as a potential predictor for various early alcohol use behaviors. Unlike other studies where distinguishing between AUD and controls or predicting alcohol initiation was more straightforward, our study tackled the challenge of predicting a more complex outcome, the trajectory of alcohol sipping behavior during development. Our objective was to uncover pivotal factors in childhood that influence the trajectories of alcohol sipping behaviors over subsequent years throughout adolescence. Since most of the participants have not had much alcohol exposure yet (i.e. full drinks), focusing on the trajectory of the number of alcohol sips over time represented by the three latent groups was appropriate in the current stage of development. In addition, our study analyzed how multilevel, multidomain factors interacted with brain structure and function at baseline and if these interactions predicted alcohol sipping patterns as children transitioned to adolescence. Our study pursued the following 3 aims: (i) to examine whether individual, family, and environmental factors measured at baseline could accurately predict patterns of alcohol sipping over time; (ii) to identify alterations in brain structure and functional connectivity at baseline that predicted alcohol sipping patterns over time; (iii) to investigate whether the multilevel multidomain factors and multimodal brain imaging features jointly and interactively predicted alcohol sipping patterns over time. We were interested in two contrasts based on our previous study: the no-sip vs. high-sip contrast, to capture an increasing trajectory of alcohol sipping over time vs. no alcohol initiation, and the low-sip vs. high-sip contrast, to differentiate between alcohol initiators with an increasing trajectory over time vs. those who had some alcohol exposure at baseline but decided to reduce their sipping over time.

## 2. Methods

### 2.1 Participants

We used the 5.1 release of The Adolescent Brain Cognitive Development (ABCD) study, the largest longitudinal study on adolescents in the United States, including a baseline cohort of 11,686 subjects coming from 9,807 unique families across 22 sites (Barch et al., 2021; Casey et al., 2018). The ABCD study aims to evaluate brain development, substance use, behavioral development, physical and mental health.

### 2.2 Measures

Multilevel, multidomain measures were selected based on a comprehensive literature review, including published primary research and review papers using search terms such as ‘adolescent alcohol use’, ‘alcohol initiation’, and ‘binge drinking’ across Google Scholar, PubMed, and the search engine for the ABCD study. For multimodal neuroimaging measures, we included all features preprocessed and made available in the ABCD study.

#### 2.2.1 Individual level

The following variables included in the current analysis were measured at the individual level at baseline and were selected based on the literature (Gu et al., 2024; Hahm et al., 2003; Hasler et al., 2015; Korhonen et al., 2009; Lopez-Caneda et al., 2014; Meque et al., 2019; Squeglia et al., 2017; Stautz & Cooper, 2013): sex assigned at birth by parent, race assigned at birth by parent, age at baseline, family history of drug use, prenatal drug exposure, body-mass index (BMI), 32 principal components of genetic ancestry, sleep time, physical activity, weekend and weekday screen time, internalizing, externalizing and conduct problems, personality traits (subscales for negative urgency, positive urgency, sensation seeking, lack of planning, and lack of perseverance; subscales for behavioral inhibitory/activation system - BIS and BAS subscales for reward responsiveness, drive, and fun seeking), child’s prosocial behavior, whether individuals have ever had a puff of tobacco or marijuana and a sip of alcohol, onset age, alcohol curiosity and intent, and positive alcohol expectancies (PAE) which was measured at 1-year follow-up due to unavailability at baseline. Additional information about measures can be found in the Supplemental materials.

#### 2.2.2 Family level

At the family level we included the following variables measured at baseline, following the literature recommendations (Hahm et al., 2003; Hallihan et al., 2023; Wells & Östberg, 2018): family income, highest parental education, parental behavior, parental monitoring, family conflict, parental rules regarding the child’s alcohol consumption and penalties for not respecting the rules, and alcohol availability in the household.

#### 2.2.3 Community level

At the community level we used the following variables in the analysis, measured at baseline, built on the rigor of prior research (Bryant et al., 2003; Huckle et al., 2010; Wells & Östberg, 2018): area deprivation index (ADI), neighborhood safety, area type, religion, academic performance, school environment, involvement and disengagement, and number of friends drinking alcohol.

#### 2.2.4 Brain structure

We used 74 cortical regions in each hemisphere parcellated by the Destrieux atlas from structural magnetic resonance imaging (sMRI) data. The ABCD sites had 3T scanners (Siemens, Phillips, GE) and participants completed 3-dimensional T1-weighted images. Details about imaging acquisition in the ABCD study can be found elsewhere (Hagler et al., 2019). We used cortical volume (VOL), including gray matter volume (GMV) and white matter volume (WMV), surface area (SA) and cortical thickness (THK) for each region of interest (ROIs) measured at baseline and we also included grey matter and white matter volume for 23 subcortical ROIs in each hemisphere derived from FreeSurfer’s automated segmentation (Fischl, 2012). We excluded participants with poor imaging quality or incomplete Freesurfer deconstruction, susceptibility to artifacts, traumatic brain injury (TBI) with loss of consciousness, or if they were referred to a clinician (Pablo Vidal-Ribas et al., 2021).

#### 2.2.5 Functional networks

Resting-state functional MRI (rs-fMRI) scans were completed at baseline and functional networks were parcellated by using the Gordon atlas(Hagler et al., 2019). We used the average pairwise ROIs correlations between and within 13 cortical networks (auditory network, audN; cingulo-opercular network, CON; cingulo-parietal network, CPN; default-mode network, DMN; dorsal attention network, DAN; fronto-parietal network, FPN; retrosplenial temporal network, RTN; salience network, SN; sensorimotor hand network, SMNh; sensorimotor mouth network, SMNm; ventral attention network, VAN; visual network, visN). We also used the average pairwise correlations between the 13 cortical networks and 10 subcortical ROIs of each hemisphere (accumbens area, amygdala, brainstem, caudate, cortex of the cerebellum, hippocampus, pallidum, putamen, thalamus proper, ventral diencephalon). We excluded participants with poor or incomplete Freesurfer deconstruction and subjects who should consider clinical referral (Pablo Vidal-Ribas et al., 2021).

#### 2.2.6 Outcome: alcohol consumption trajectories

To capture the alcohol use behavior in children as they transition into adolescence, we previously identified three latent longitudinal patterns of alcohol sipping in 9–10-year-old adolescents from the ABCD study: early low-sip alcohol initiation, early high-sip alcohol initiation, and no alcohol initiation (Fig. 1) (Ferariu et al., 2024). Given that alcohol sipping in children is highly heterogeneous, identifying latent trajectories allowed us to uncover distinct subtypes of early alcohol exposure, providing a more detailed understanding of who could be at greater risk for continued alcohol use versus those who would more likely refrain. Among the participants exhibiting early alcohol experimentation, our approach further differentiated between those who continued their alcohol use (i.e., high-sip group) vs. those who started but then reduced their alcohol use (i.e., low-sip group). In the current study, we focused on two contrasts as our categorical outcome variables: no-sip vs. high-sip and low-sip vs. high-sip. We excluded the no-sip vs. low-sip contrast, because we were particularly interested in the prediction of (i) those who increased their sipping over time (high-sip group) vs. those who have not yet initiated alcohol sipping (no-sip group) and (ii) among individuals who initiated alcohol sipping early, those who continued sipping over time (high-sip group) vs. those who reduced their sipping over time (low-sip group). Fig. 1 shows the average curve for the number of alcohol sips over time for the three groups: no-sip (84.22%, N = 9700), low-sip (5.34%, N = 615) and high-sip (10.44%, N = 1202).

**Fig. 1.**
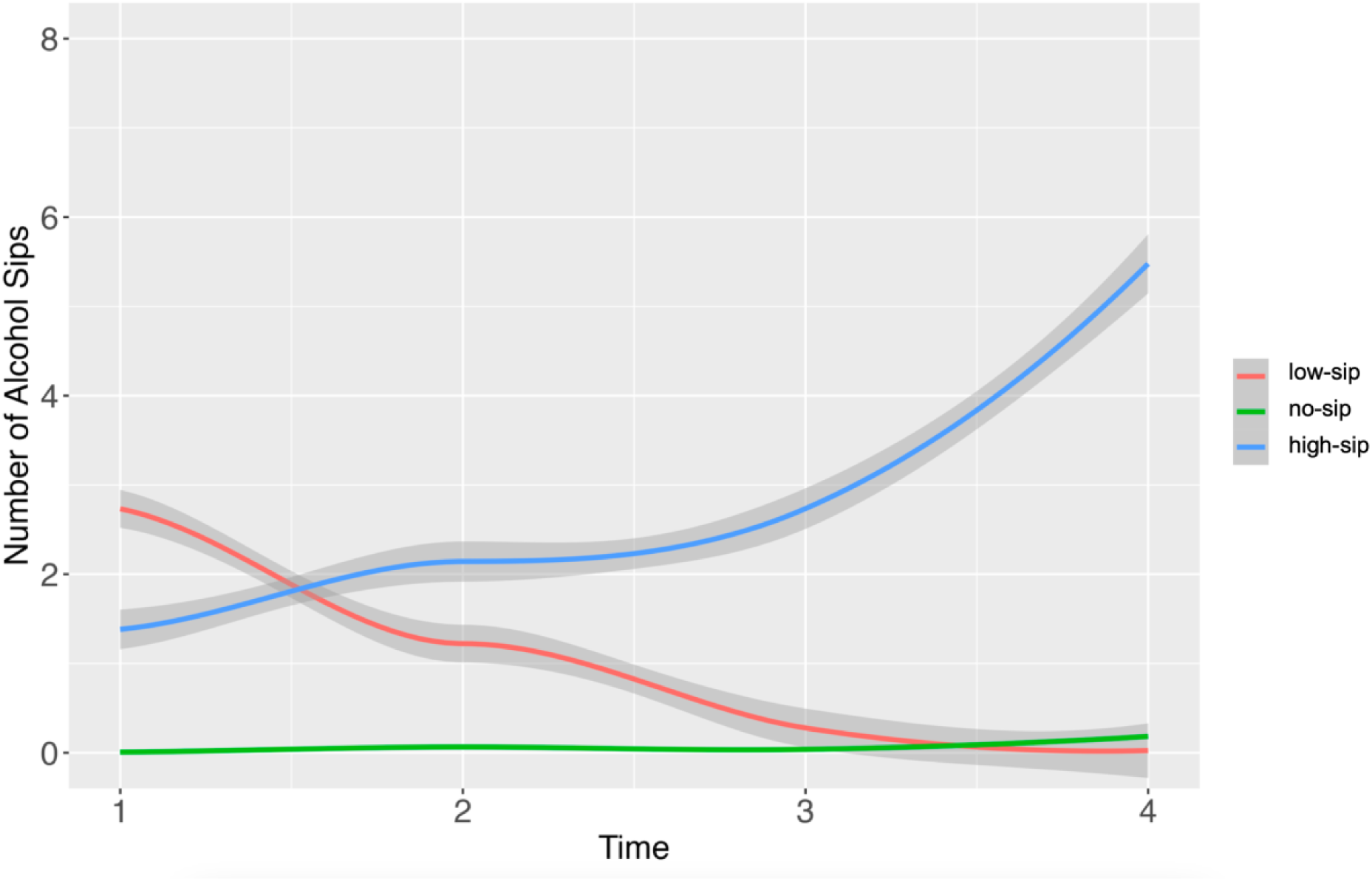
The average curve for the latent classes representing the sipping behavior over time in the ABCD cohort and the corresponding standard error.

### 2.3 Statistical Analysis

We considered a data-driven approach to identify all of the relevant multilevel multidomain factors and brain imaging features for predicting alcohol sipping patterns. The advantages of using a data-driven approach over a hypothesis-driven approach include the possibility of examining a large number of factors and their interactions, and the property of allowing the data to inform us which multilevel multidomain factors and brain imaging features are relevant and of most importance in predicting alcohol sipping patterns as children transition into adulthood.

#### 2.3.1 Aim 1

In Aim 1, we evaluated and compared several popular machine learning methods, such as ensemble learning (Lappalainen & Miskin, 2000), classification and regression trees (CART) (Breiman, 2017), random forest (Breiman, 2001), support vector machines (SVM) (Smola & Schölkopf, 2004) and regularized regression (Ridge, LASSO, Elastic Net) (Hoerl & Kennard, 1970; Tibshirani, 1996; Zou & Hastie, 2005) to classify the membership of the alcohol sipping variable (no-sip vs. high-sip and low-sip vs. high-sip) by examining the multilevel, multidomain factors. We split the data into a training set (75% of the data) and a test set (the remaining 25%). During the training stage, we used a 10-fold cross-validation (CV) to split the training data into a sub-training set and a validation set. For the sub-training set, we implemented resampling-based strategy to over- and under-sample the majority and minority groups, respectively, since our data was unbalanced (no-sip:low-sip:high-sip ratio of 16:1:8) (He & Garcia, 2009). Moreover, we used validation sets from the 10-fold CV to tune the model parameters for each of the methods described above. To assess model fit, we evaluated area under the receiver operating characteristic (ROC) curve (AUC), sensitivity and specificity. By using machine learning methods, we expected to identify the most important features that could accurately predict alcohol sipping patterns in adolescents. Specifically, we tested whether the models predicted the outcomes significantly above the chance level, as indicated by a 95% confidence interval of AUC not including 0.50.

#### 2.3.2 Aim 2

In Aim 2, we used multivariate pattern analysis (MVPA) to better understand the complex relationships between brain structure, functional networks, and alcohol sipping behavior. We repeated the statistical analysis in Aim 1, with each imaging modality (sMRI and rs-FMRI) used as input data separately and then combined. For the structural features, we trained the models on each type of structural feature (volume, cortical thickness, and surface area) individually and then also combined. We included sex, age at baseline, family history of drug use and SES as covariates.

#### 2.3.3 Aim 3

In Aim 3, we conducted moderation analysis to investigate how the top predictive multilevel, multidomain factors identified in Aim 1 interact with the top predictive brain structures and functional networks identified in Aim 2 in the classification of the alcohol sipping groups. We utilized iterative random forest (iRF) for its ability to uncover complex interactions among variables predicting alcohol sipping patterns in adolescents (no-sip vs. high-sip and low-sip vs. high-sip) (Basu et al., 2018). The iRF method enhances traditional random forest (RF) algorithms by iteratively refining feature importance weights based on previous RF runs. It stabilizes these weights by bootstrapping samples, allowing it to identify co-occurring features that are representative of alcohol sipping classes. The iRF was trained, validated and tested similarly to the other machine learning techniques discussed in Aim 1. We used two additional tuning parameters obtained through CV, 500 bootstrap samples and 10 iterations.

#### 2.3.4 Feature Selection

We performed feature selection to identify the top predictive features since machine learning models do not generate p-values. We derived feature importance scores (FIS) to rank features from the most important to the least important based on how much they contribute to the prediction of the outcome. Where feature selection was not directly performed by the machine learning models, feature importance was assessed using recursive feature elimination (RFE), which computed the ratio between the AUC of the full model (including all variables) and the AUC of reduced models, each omitting one variable. An importance score ratio greater than 1 indicates that the eliminated feature is significant for predicting the outcome. The top predictive features were selected if their importance scores exceeded the mean by one standard deviation. Due to the black box nature of machine learning models, it is not possible to obtain coefficients or p-values that represent the effect of each individual factor, therefore we used accumulated local effects plot (ALE) to determine the directionality of the predictor on the outcome (Apley & Zhu, 2020). ALE plots illustrate how different values within a feature’s range influence the average prediction of a machine learning model. These accumulated effects are centered at zero, making it possible to interpret how the model’s predictions shift depending on whether the effect is positive or negative. For instance, ALE values greater than zero represent positive prediction values in comparison with the average prediction.

## 3. Results

### 3.1 Aim 1: Multilevel, Multidomain Factors Predicting Alcohol Sipping Patterns

#### 3.1.1 No-vs-High sip Contrast

Table S1 presents the performance metrics for all models using multilevel, multidomain variables as input for the no-vs-high sip contrast. The Ridge regression model demonstrated the best performance, achieving an AUC of 0.707 (95% CI: 0.674, 0.740), with sensitivity of 0.646 and specificity of 0.768, significantly above chance level. Feature importance was assessed using recursive feature elimination (RFE), and all variables exceeded the threshold of importance score ratio of 1, underscoring their relevance in predicting the no-vs-high sip contrast. Fig. 2 shows the ALE plots of the features with importance scores (FIS) exceeding the mean importance score by one standard deviation. Participants with positive alcohol expectancies (PAE, FIS=1.038, ALE in Fig 2j) and those who expressed their curiosity (FIS= 1.043, ALE in Fig 2d) or intent (FIS = 1.042, ALE in Fig 2g) to try alcohol were more likely to be classified into the high-sip group. Higher values of parental monitoring (FIS = 1.042, ALE in Fig 2e) and longer screentime during weekends (FIS = 1.051, ALE in Fig 2a), along with individuals engaging in more sports activities (FIS = 1.042, ALE in Fig 2f) and coming from a better school environment (FIS = 1.040, ALE in Fig 2h) were also associated with this classification.

**Fig 2.**
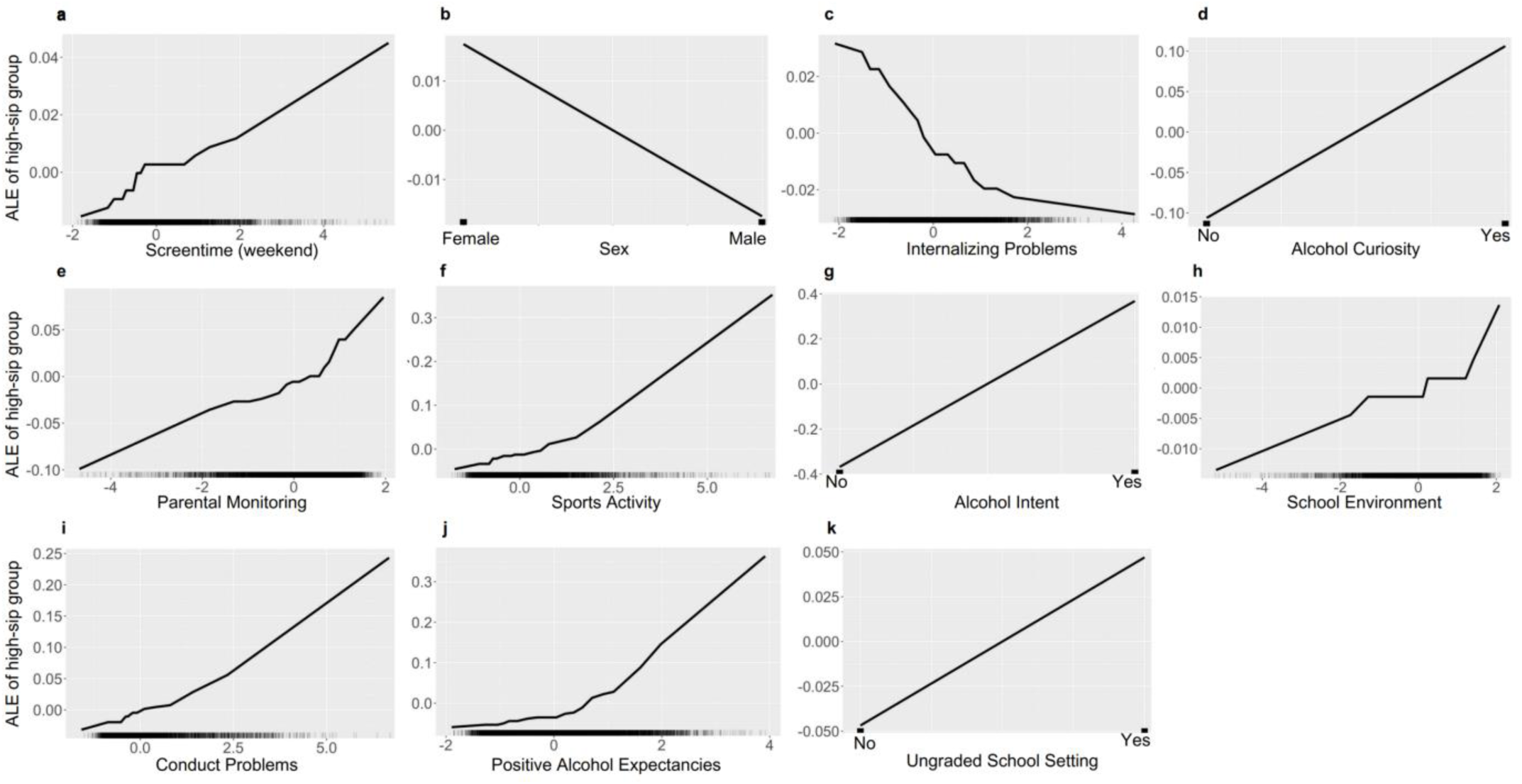
Accumulated local effects (ALE) plots of main effects in the classification of no-sip vs. high-sip based on the Ridge model.

Moreover, participants having ungraded school settings (FIS = 1.038, ALE in Fig 2k), as well as participants with more conduct problems (FIS = 1.039, ALE in Fig 2i) were also classified into the high-sip group. Participants with ungraded school setting were either not in school or their school system didn’t provide grades. Conversely, participants with more internalizing problems (FIS = 1.044, ALE in Fig 2c) and male individuals (FIS = 1.046, ALE in Fig 2b) were more likely to be classified in the no-sip group.

#### 3.1.2 Low-vs-High sip Contrast

Table S2 shows the performance metrics for all models using multilevel, multidomain variables as input for the low-vs-high-sip contrast. The ensemble learning model which included linear SVM, Ridge regression, and CART achieved the best performance significantly better than chance level, with AUC of 0.590 (95% CI: 0.536, 0.645), sensitivity of 0.404 and specificity of 0.777. Feature importance derived by the algorithm of the ensemble model using the overall score and was calculated based on all the individual models (Kuhn, 2008). ALE plots shown in Fig. 3 were used to assess the directionality of predictions for each feature whose FIS exceeded the mean plus one standard deviation. Individuals with higher levels of the 21^st^ principal component of genetic ancestry (FIS = 2.569, ALE in Fig 3a), parental monitoring (FIS = 1.951, ALE in Fig 3c), family conflict (FIS = 1.361, ALE in Fig 3h), and better school environment (FIS = 1.242, ALE in Fig 3i), along with older participants (FIS = 1.686, ALE in Fig 3e), were more likely to be classified in the high-sip group. In contrast, individuals experiencing more childhood trauma (FIS = 1.429, ALE in Fig 3g), coming from areas with increased ADI (FIS = 1.803, ALE in Fig 3d), having greater levels of the 23^rd^ principal component of genetic ancestry (FIS = 1.588, ALE in Fig 3f), and being male (FIS = 2.162, ALE in Fig 3b) were less likely to be classified in the high-sip group. Additional features with weaker effects on the outcome included: school involvement, the 6^th^ principal component of genetic ancestry, lack of perseverance, lack of planning, parental behavior and sports activity.

**Fig. 3.**
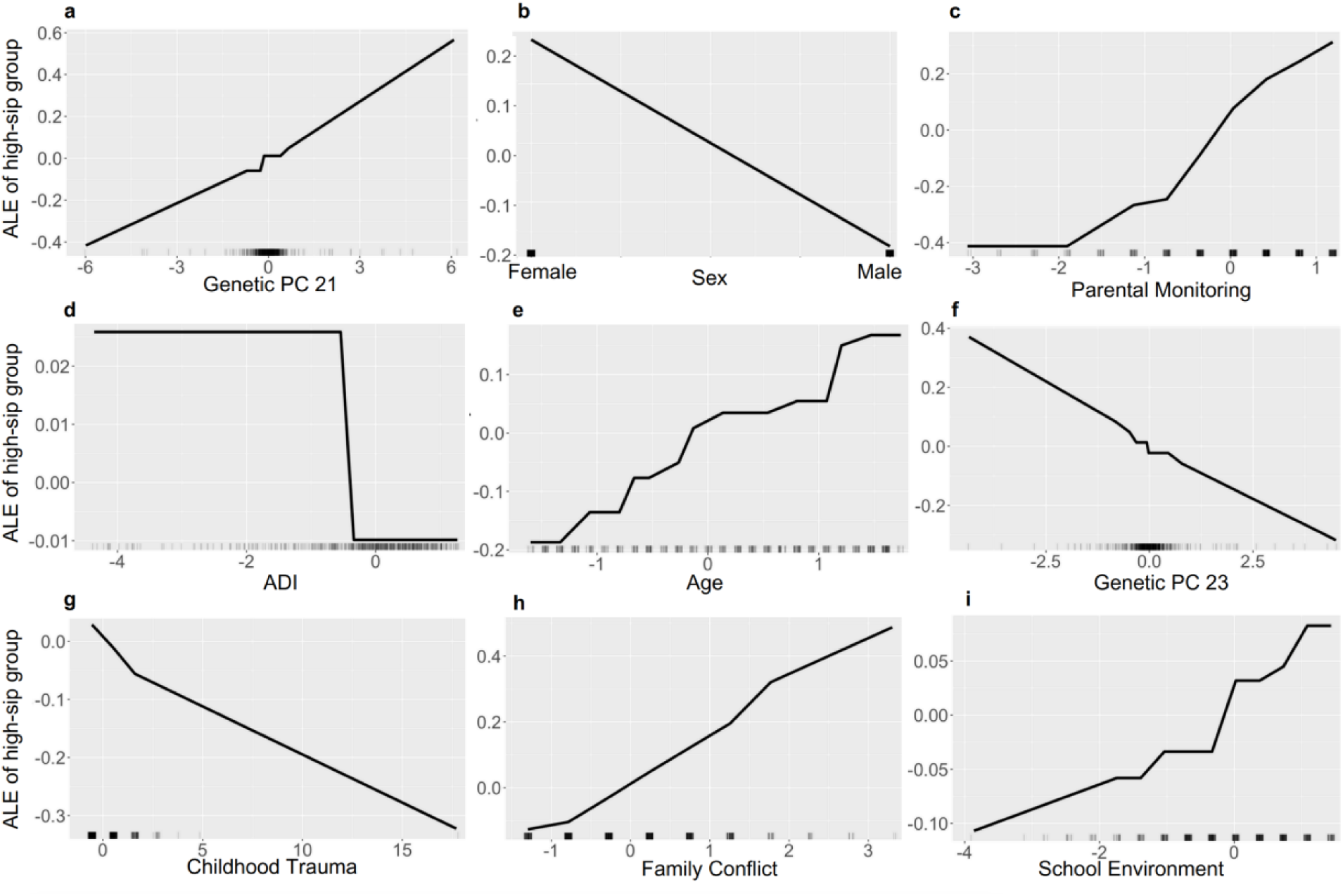
Accumulated local effects (ALE) plots of main effects in the classification of low-sip vs. high-sip based on the ensemble learning model.

### 3.2 Aim 2: Multimodal Neuroimaging Features Predicting Alcohol Sipping Patterns

#### 3.2.1 No-vs-High sip Contrast

Table S3 contains the performances of all models using structural features and functional connectivity as input data. Using multimodalities in the prediction of the no-vs-high-sip groups was more informative than using single modalities. The best performing model with neuroimaging features was the Elastic Net regression, showing an AUC of 0.621 (95% CI: 0.589, 0.652), sensitivity of 0.651 and specificity of 0.591, demonstrating performance significantly above chance level. The ALE plots of the sMRI features and rs-FC features, extracted based on the non-zero coefficients from the Elastic net model are shown in Fig. 4 and Fig. 5, respectively. Older participants (b=0.126, ALE in Fig 4a), along with participants showing greater cortical volume in the right anterior transverse collateral sulcus (b=0.047, ALE in Fig 4b) were more likely to be classified in the high-sip group. Moreover, increased functional connectivity between SMN hand and right nucleus accumbens (b=0.042, ALE in Fig 5a)., between CON and left amygdala (b=0.031, ALE in Fig 5b) or right hippocampus (b=0.022, ALE in Fig 5c), and between visual network and left hippocampus (b=0.014, ALE in Fig 5d) or right pallidum (b=0.006, ALE in Fig 5e) were predictive of the high-sip group. Conversely, greater cortical thickness in the right lateral orbital sulcus (b=-0.011, ALE in Fig 4c), left horizontal ramus of the anterior segment of the lateral sulcus (b=-0.029, ALE in Fig 4e) and right transverse frontopolar gyri and sulci (b=-0.057, ALE in Fig 4f), along with increased functional connectivity between auditory network and left putamen (b=-0.012, ALE in Fig 5f) were predictive of the no-sip group. Participants with higher ADI (b=-0.108, ALE in Fig 4d) were also classified into the no-sip group. Additional features with weaker effect on the outcome included: surface area in the right inferior temporal sulcus, cortical volume in the right middle frontal gyrus, cortical thickness in the inferior frontal gyrus operculum, cortical thickness in the left intraparietal sulcus and transverse parietal sulci, functional connectivity between DMN and right amygdala or left nucleus accumbens, and between SMN hand and SMN mouth.

**Fig. 4.**
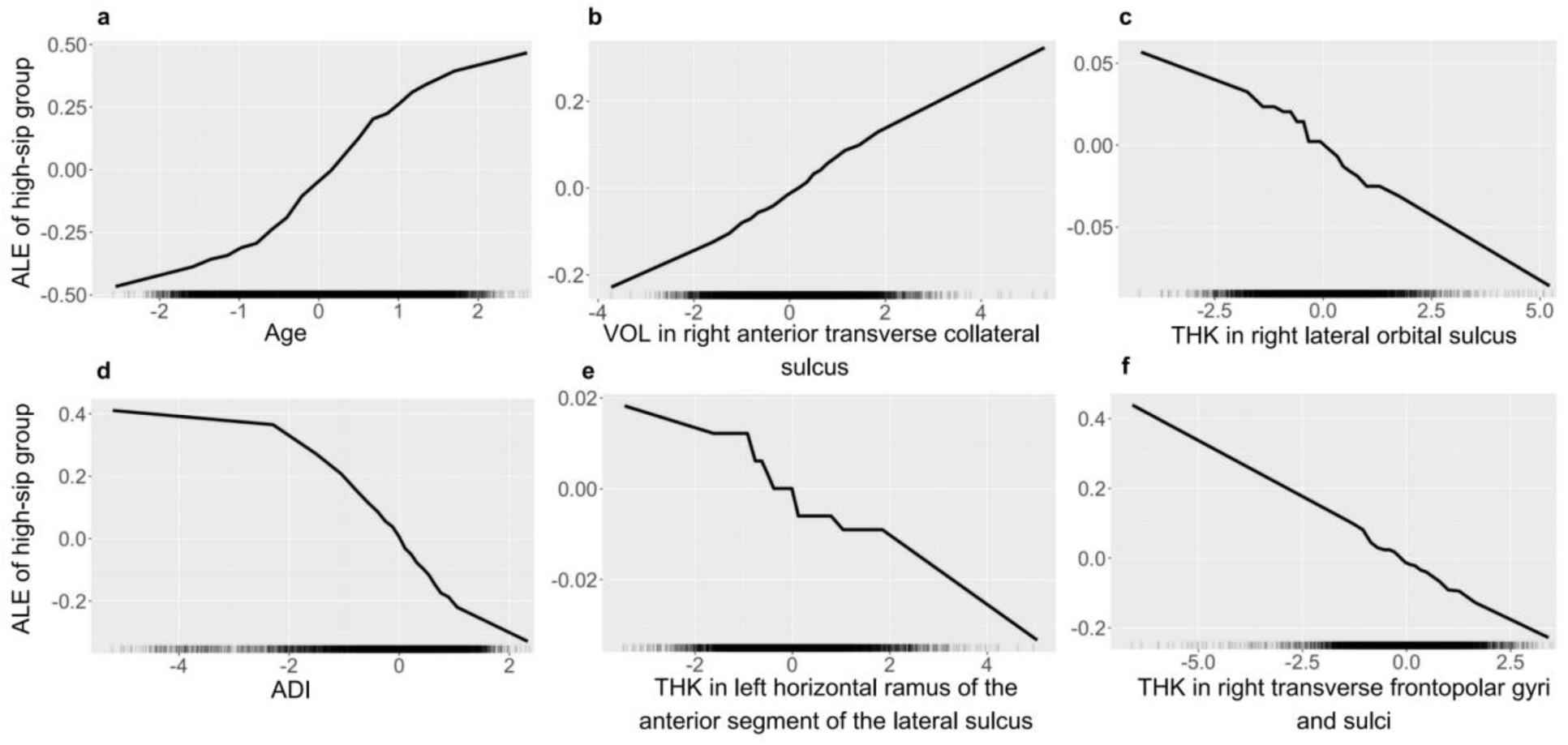
Accumulated local effects (ALE) plots of demographics and structural features in the classification of no-sip vs. high-sip based on the Elastic net model.

**Fig. 5.**
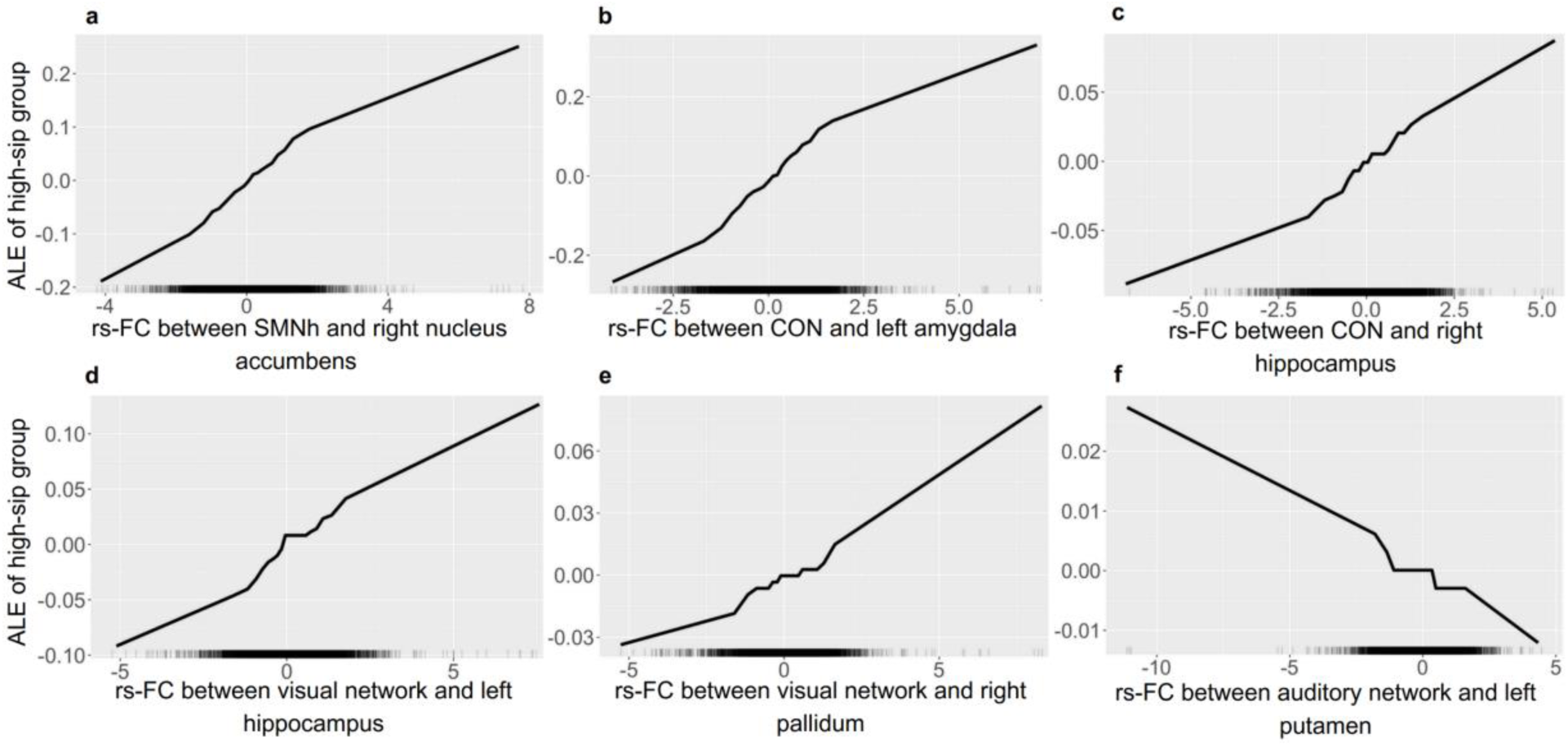
Accumulated local effects (ALE) plots of functional connectivity features in the classification of no-sip vs. high-sip based on the Elastic net model.

#### 3.2.2 Low-vs-high sip Contrast

Functional connectivity was more informative in the prediction of low-vs-high-sip groups compared to sMRI features alone or to both modalities combined. Table S4 shows the performances of the models with functional connectivity as input data in the classification of the low-vs-high sip contrast. The best performing model was an Elastic Net model achieving an AUC of 0.560 (95% CI: 0.506, 0.613), sensitivity of 0.530 and specificity of 0.589, demonstrating performance significantly above chance level. Fig. S1 shows the ALE plots for the features with non-zero coefficients extracted from the Elastic Net model. Older participants (b=0.058, ALE in Fig S1t), along with participants showing greater functional connectivity between visual network and right nucleus accumbens (b=0.002, ALE in Fig S1g) or DAN (b=0.009, ALE in Fig S1h), between CON and right ventral diencephalon (b=0.002, ALE in Fig S1i), between none network and right nucleus accumbens (b=0.021, ALE in Fig S1j), between DAN and left amygdala (b=0.023, ALE in Fig S1k) or visual network (b=0.035, ALE in Fig S1l), between DMN and right putamen (b=0.023, ALE in Fig S1m), between RTN and auditory network (b=0.032, ALE in Fig S1n) or left ventral diencephalon (b=0.074, ALE in Fig S1o), between VAN and RTN (b=0.044, ALE in Fig S1s), between SN and the left cortex of the cerebellum (b=0.048, ALE in Fig S1p), between auditory network and RTN (b=0.050, ALE in Fig S1r) and between FPN and right caudate (b=0.102, ALE in Fig S1q) were more likely to be classified into the high-sip group. Participants less likely to be categorized into the high-sip group were males (b=-0.103, ALE in Fig S1a), coming from greater ADI areas (b=-0.074, ALE in Fig S1b) and showed increased functional connectivity between auditory network and right amygdala (b=-0.037, ALE in Fig S1c), between SN and DMN (b=-0.037, ALE in Fig S1d) and between SMN mouth and right amygdala (b=-0.017, ALE in Fig S1e) or right cortex of the cerebellum (b=-0.012, ALE in Fig S1f). Additional features with weaker effect on the outcome included: functional connectivity between visual network and FPN, between FPN and left amygdala or right nucleus accumbens, between DMN and right cortex of the cerebellum, between CON and SN or left pallidum or right caudate, between RTN and left cortex of the cerebellum, between none network and left pallidum, between VAN and right thalamus proper, between auditory network and VAN, between DAN and left ventral diencephalon, between SMN mouth and right pallidum and between SMN hand and left nucleus accumbens.

### 3.3 Aim 3: The Relationship between Multilevel, Multidomain and Multimodal Neuroimaging Features in the Prediction of Alcohol Sipping Patterns

#### 3.3.1 No-vs-High sip Contrast

The model performance of the iRF model for the no-vs-high sip contrast achieved performance above chance level, showing an AUC of 0.704 (95% CI: 0.670, 0.739), sensitivity of 0.605 and specificity of 0.804. Twenty-seven interactions with stability scores greater than 0.50 were identified, and those with stability greater than 0.75 are visualized in Fig. 6. Of the top 9 stable interactions, only second-order interactions were identified. The ALE plots for main effects and interaction effects are shown in Fig. 7 and Fig S1, respectively.

**Fig. 6.**
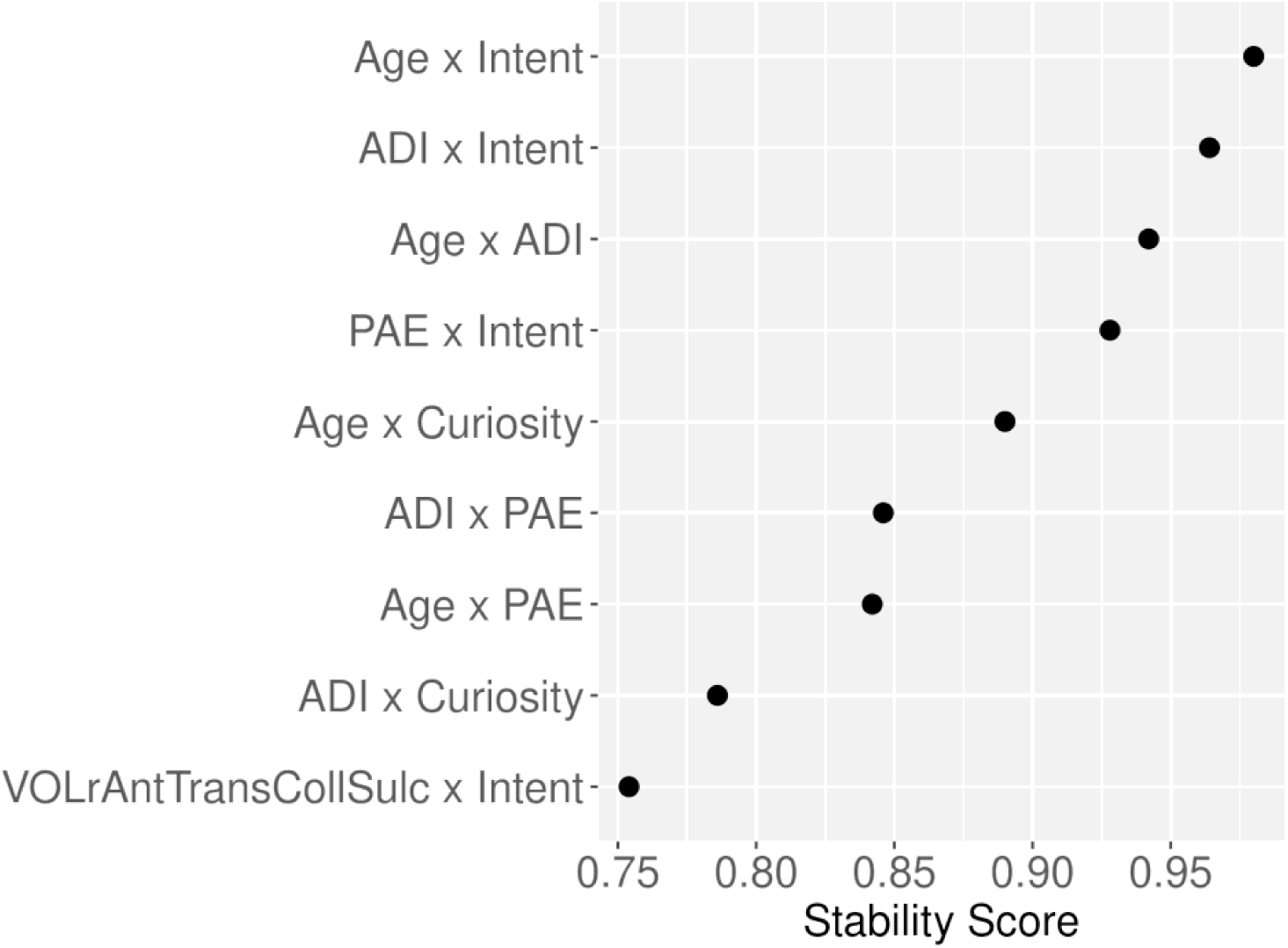
Interactions recovered by iRF with stability scores greater than 0.75, for the no-sip vs. high-sip contrast. *Note: ADI – area deprivation index, PAE – positive alcohol expectancies, VOLrAntTransCollSulc – cortical volume in the right anterior transverse collateral sulcus*.

**Fig. 7.**
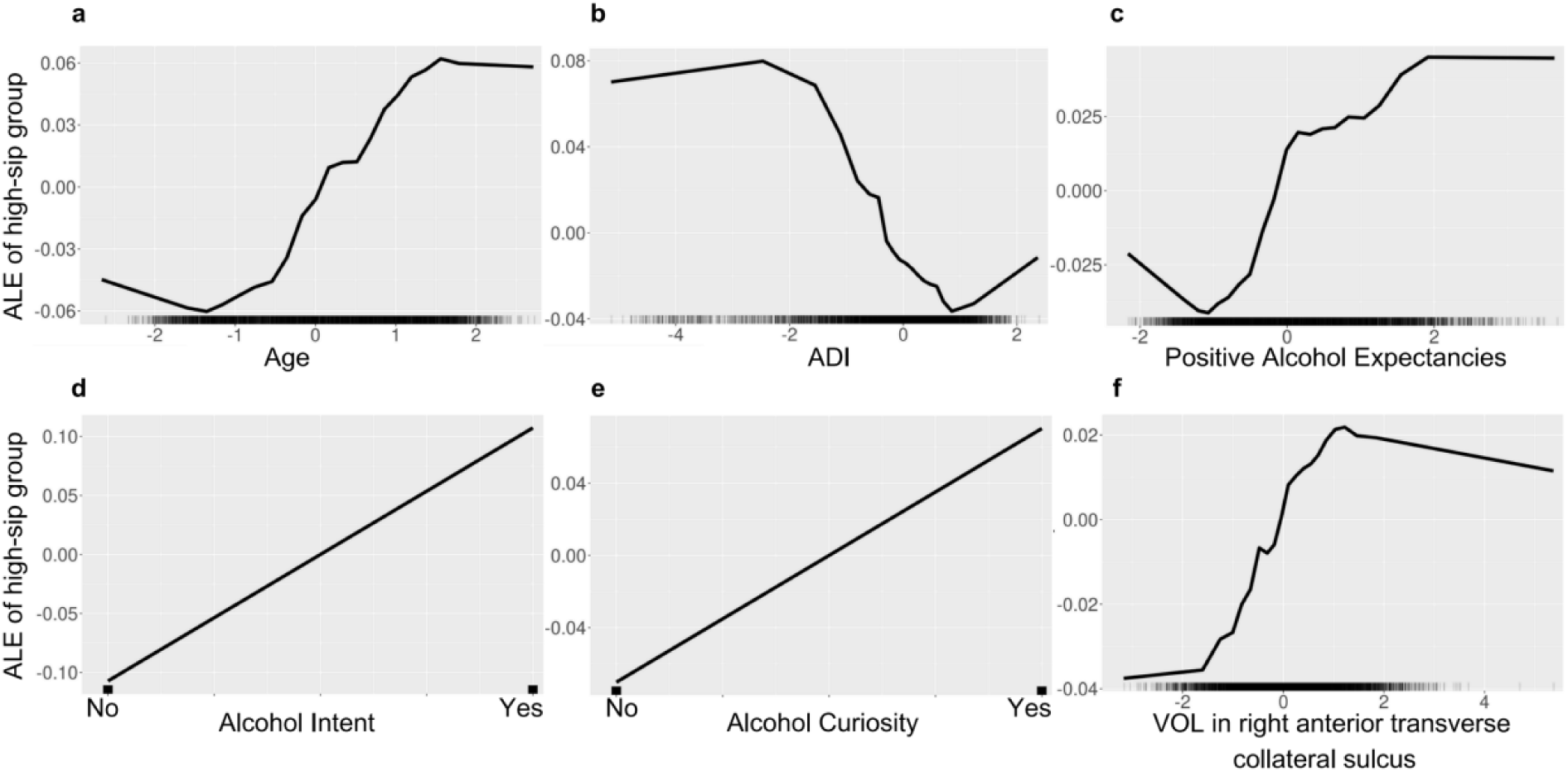
Accumulated local effects (ALE) plots of main effects in the classification of no-sip vs. high-sip based on the iRF model.

Interestingly, older participants (Fig 7a), or those with higher PAE (Fig 7c), having the intent (Fig 7d) or curiosity (Fig 7e) to try alcohol, as well as participants with greater cortical volume in the right anterior transverse collateral sulcus (Fig 7f) were more likely to be in the high-sip group. We also found a stable interaction between age and alcohol intent, showing an additional increase in the likelihood of belonging to the high-sip group for younger participants having the intent to try alcohol (Fig S2a), above the individual effects of age and alcohol intent. This combined effect of age and alcohol intent on the high-sip classification is greater than the sum of their main effects. Having the intent to try alcohol at baseline, as well as showing greater cortical volume in the right anterior transverse collateral sulcus (Fig S2c) or having greater PAE (Fig S2d) similarly showed an increased likelihood of high-sip classification, after accounting for main effects. Low ADI showed an increased effect in the likelihood of belonging to the high-sip group, but this effect decreased as ADI increased (Fig 7b). In the interaction with age, younger participants coming from slightly lower ADI were less likely to belong to the high-sip group, but as age increased, the likelihood for the high-sip group increased within the same ADI interval (Fig S2g). Participants coming from low ADI, even though showing low PAE (Fig S2e) or no intent (Fig S2b) or no curiosity (Fig S2i) to try alcohol, had an additional increased effect on the likelihood to belong to the high-sip group, after accounting for the main effects. Older participants with greater PAE (Fig S2f) or those who were curious to try alcohol (Fig S2h) were also more likely to belong to the high-sip group, after accounting for main effects.

#### 3.3.2 Low-vs-High sip Contrast

The iRF model for the low-vs-high sip contrast performed significantly above chance level, achieving an AUC of 0.593 (95% CI: 0.536, 0.650), sensitivity of 0.700 and specificity of 0.486. Additionally, there were 12 interactions with stability scores above 0.50, and those with stability greater than 0.75 are visualized in Fig. 8. The ALE plots for the main effects and interaction terms are shown in Fig. 8 and Fig. S3, respectively.

**Fig. 8.**
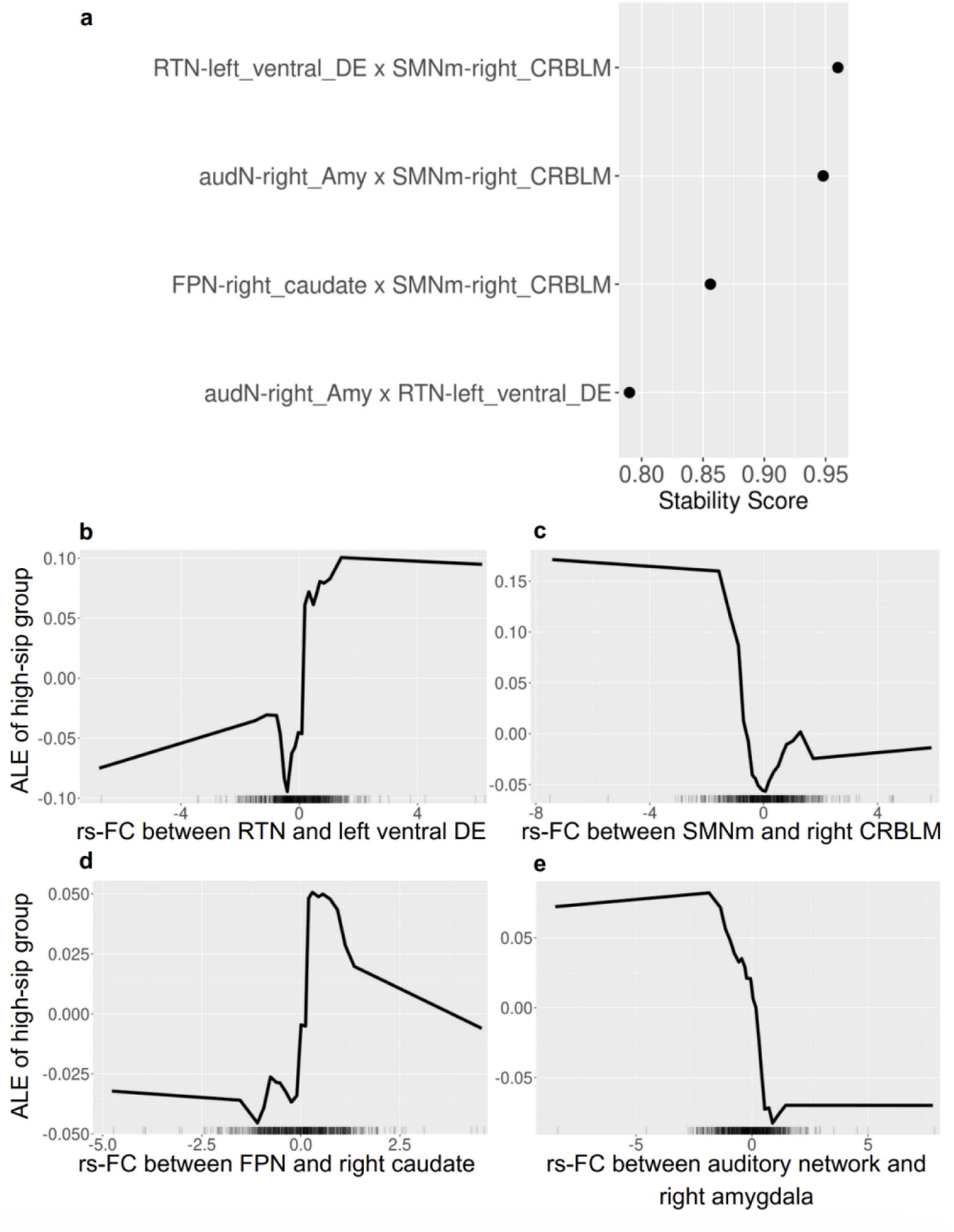
(a) Interactions recovered by iRF with stability scores greater than 0.75, for the low-sip vs. high-sip contrast; (b-e) Accumulated local effects (ALE) plots of main effects in the classification of low-sip vs. high-sip based on the iRF model. *Note: RTN - retrosplenial temporal network, CRBLM – cerebellum cortex, audN – auditory network, SMNm - sensorimotor mouth network, DE – diencephalon, Amy – amygdala*.

Participants with greater rs-FC between RTN and left ventral diencephalon (Fig 8b), and between FPN and right caudate (Fig 8d), along with participants with lower rs-FC between SMN mouth and right cerebellum (Fig 8c) and between auditory network and right amygdala (Fig 8e) were more likely to belong to the high-sip group. We also found stable interactions between rs-FC among these networks. Having lower rs-FC between RTN and left ventral DE, as well as presenting lower rs-FC between SMN mouth and right cerebellum showed an additional increase in the likelihood of belonging to the high-sip group (Fig S3a), above the individual effects of rs-FC between RTN and left ventral DE and of rs-FC between SMN mouth and right cerebellum separately. Showing hyperconnectivity between auditory network and right amygdala, as well as having hypoconnectivity between SMN mouth and right cerebellum (Fig S3b) or between RTN and left ventral diencephalon (Fig S3d) also showed an increased likelihood of high-sip classification, after accounting for main effects. Similar classification was shown for participants with hyperconnectivity between FPN and right caudate and hypoconnectivity between SMN mouth and right cerebellum (Fig S3c), after accounting for the main effects.

## 4. Discussion

This study conducted a comprehensive analysis to identify key features predictive of alcohol sipping patterns in children as they transition to adolescence. Using popular machine learning models and baseline data from the ABCD study, we extracted features that distinguish between early drinking onset and a more stable, non-sipping pattern over time, as expected in such a young cohort. Among children who began sipping alcohol at a younger age, we further explored the features that differentiate those who continued sipping from those who reduced their alcohol consumption over time. While previous studies have often limited their analysis to specific types of input data, such as focusing solely on brain structure or function, or emphasizing individual versus environmental factors, our research fills a critical gap in the literature by examining multilevel, multidomain and multimodal neuroimaging factors together.

We proposed and achieved three aims where we analyzed various factors in the prediction of previously identified alcohol sipping patterns over time. In Aims 1 and 2 we used multilevel, multidomain features and multimodal neuroimaging features, respectively, as input variables in the machine learning models. In Aim 3 we included the most predictive features from the previous aims, as well as their interactions. We found that, among individuals who experimented with alcohol at baseline, functional connectivity features, along with their interactions, distinguished between those who continued sipping and those who chose to reduce their sipping. Moreover, this classification was also supported by factors related to the individual (sex, age, component of genetic ancestry), related to the family (parental monitoring, family conflict, childhood trauma), and related to the environment (ADI, school environment). Additionally, both structural and functional features differentiated between those with an increasing sipping trajectory over time and those who abstained from further alcohol experimentation. Factors related to the individual (sex, conduct and internalizing problems, lifestyle factors, attitudes towards drinking), family (parental monitoring), and environment (school environment, grades) were found to be predictive of this classification. Moreover, the brain structural features combined with age and interactions involving positive alcohol expectancies, ADI, curiosity and intent to try alcohol also supported this differentiation. Therefore, we showed that multilevel multidomain factors and brain structural and functional connectivity features can predict the pattern of alcohol sipping over time in adolescence.

The machine learning models used in our analysis achieved performance levels (AUCs ranging from 0.56 to 0.71) comparable to similar studies on alcohol initiation, despite differences in input data. For instance, research using the ABCD dataset to predict alcohol initiation based on contextual factors, inhibitory control, and reward sensitivity reported AUCs ranging from 0.59 to 0.67 (Moore et al., 2024). Another study investigating alcohol use frequency among 14-to 16-year-old adolescents achieved an AUC of 0.74 using only behavioral data (Amialchuk et al., 2021). In adult studies, machine learning models demonstrated AUCs between 0.70 and 0.80 for tasks like classifying individuals with AUD (Lee et al., 2019; Shukla et al., 2018) or identifying unhealthy drinking during early adulthood (Bi et al., 2013; Gowin et al., 2021). However, these tasks captured only a snapshot of alcohol use behavior rather than trajectories of alcohol sipping patterns over time in healthy children and they utilized only subsets of the broad range of features included in our analysis.

Previous research has identified several multilevel, multidomain features as predictive of alcohol initiation and binge drinking in older adolescents. At the individual level, positive attitudes towards drinking have been associated with binge drinking and severe alcohol use (DiBello et al., 2018). On the contrary, negative attitudes towards drinking have been associated with reduced alcohol consumption (Hahm et al., 2003). In our study, increased positive alcohol expectancies and having the curiosity or intent to try alcohol distinguished those who increasingly sipped alcohol over time (high-sip group) from those who had not yet initiated alcohol sipping (no-sip group). Additionally, prior research has shown that SES significantly influences alcohol consumption, with individuals from higher SES backgrounds tending to drink more frequently (Huckle et al., 2010). We found stable interactions between age, ADI, positive alcohol expectancies, curiosity and intent to try alcohol in the prediction of the high-sip group. Being younger or having greater positive alcohol expectancies along with having the intent to try alcohol would increase the likelihood of the participants to engage in more alcohol sipping over time. Coming from a neighborhood with lower ADI, equivalent to higher SES, even though having no intent or curiosity to try alcohol, having lower positive alcohol expectancies or being older, would add an additional positive effect in the prediction of belonging to the high-sip group, compared to the average prediction. These findings are consistent with the Social-Ecological Model and highlight the complex interplay between risk factors, such as attitudes towards drinking, age, and environment, which can interact with each other by amplifying or buffering the effect on the development of early alcohol use patterns.

Another interesting finding in our study was that family conflict, parental monitoring, along with a better school environment and a sedentary lifestyle, reflected by increased screentime during weekends, appeared as important features predicting the high-sip group. High levels of family conflict, along with parental monitoring and behavior, are commonly associated with the development and maintenance of anxiety disorders in children and socially anxious youth may be more prone to engage in alcohol use or develop alcohol-related problems (Blumenthal et al., 2010; Ginsburg et al., 2018). Our findings could indicate that this type of family and school environment may play a significant role in influencing decisions to engage in higher levels of alcohol consumption over time.

Although previous research has identified childhood trauma, conduct problems, and internalizing problems as risk factors for high-risk behaviors, such as binge drinking (Aloi et al., 2020; Meque et al., 2019), our findings are only partially consistent with the existing literature. Greater levels of conduct problems classified the participants in our study into the high-sip group, while greater internalizing problems and childhood trauma levels were predictive of no-sip group and low-sip group, respectively. Given that all of the factors were measured at baseline, it is possible that participants experiencing childhood trauma had an increased experience with alcohol initially but then decreased their consumption over time. This pattern suggests that while childhood trauma is a well-established risk factor for AUD onset, its influence on early alcohol use behaviors might be more complex and influenced by other factors, such as social environment, coping mechanisms and resilience (Enoch, 2011; Wingo et al., 2014). Nevertheless, our findings suggest that these factors may become problematic in alcohol experimentation if they persist over time, and it is important to track the participants at risk in the future.

AUD is a complex genetic disorder, where genetic ancestry has been found to be associated with problematic alcohol use behaviors (Zhou et al., 2023). Principal components of genetic ancestry represent underlying population structure and genetic variation, capturing the major axes of genetic differentiation across individuals. Our analysis revealed two principal components of genetic ancestry as predictors of the group with the increasing sipping trajectory over time versus the one with a decreasing trajectory. To our knowledge, this is one of the first studies including genetic ancestry in the relationship with alcohol sipping patterns in the ABCD cohort. These findings could add valuable information about how genetic ancestry can impact alcohol sipping, and how certain populations are at risk for developing AUD.

Extensive research has examined the brain structures and functions disrupted by alcohol use but predicting which individuals will develop problematic drinking and AUD based on brain alterations is more challenging. This difficulty arises because alcohol-related changes affect development, which progresses at different rates for individuals, with additional variations due to sex and other factors. Honarvar et al. (2023) conducted the first systematic review on neuroanatomical predictors of problematic alcohol use in adolescents, and as expected, the findings were heterogeneous. Generally, thinner cortices in the frontal (including dlPFC, IFG and frontal pole), parietal, and temporal (including ITC) regions were linked to alcohol initiation or problematic use in older adolescents (Honarvar et al., 2023). We found that increased cortical thickness in adjacent regions to the right IFG, specifically in the right lateral orbital sulcus, as well as in the right transverse frontopolar gyri and sulci and in the left anterior segment of the Sylvian fissure, was predictive of the no-sip group in the ABCD cohort of 9-10-year-olds. Therefore, thinner cortices in these areas would be predictive of the high-sip group. Previous studies on older adolescents have found that smaller gray matter volume in the frontal and temporal regions may be a predisposing risk factor for future alcohol use and binge drinking (Baranger et al., 2020; Robert et al., 2020). However, our study found that increased cortical volume in the right anterior transverse collateral sulcus was predictive of the high-sip group, but it is crucial to emphasize the importance of neurodevelopment in our findings. At such a young age, 9-10 years old, the increase of cortical volume in these frontal and temporal regions might reflect normal development rather than a biomarker of increasing alcohol sipping over time. Our iRF model detected stable interactions between the intent to try alcohol and brain structure predicting the high-sip group. Having the intention to try alcohol soon and having increased cortical volume in the right anterior traverse collateral sulcus would increase the likelihood of the prediction of the high-sip group, compared to the average prediction. This indicates that relying solely on brain structure metrics may not adequately predict which children will increase their alcohol use over time, but we must consider how structural features change during development and how they might be impacted by other factors.

As far as we know, whole-brain FC analysis using fMRI has not been performed on adolescents as young as 9-10 years old in the prediction of their future alcohol behavior. To fill this gap, we included cortical and subcortical functional networks in the prediction of alcohol sipping patterns. In predicting participants who developed an increasing pattern of alcohol sipping over time, we identified several network connectivities as important features. Increased rs-FC between the SN, DMN, or FPN and subcortical regions related to reward and emotion was predictive of those with an increasing alcohol sipping trajectory over time. These findings are in line with DeWitt et al. (2014), who also found increased FC in emotion, reward, executive control, and default mode networks to be associated with risk-taking behavior, such as binge-drinking, in older adolescents (DeWitt et al., 2014). Similarly, Morales et al. (2021) reported increased rs-FC between the DMN and ventral-striatal regions, to be linked with earlier binge drinking behavior (Morales et al., 2021).

In alcohol-dependent patients, Seo et al. (2013) found that increased FC between the ventromedial prefrontal cortex (vmPFC), a key region in the DMN, and the anterior cingulate cortex (ACC), a key region of the SN, was correlated with alcohol cravings during early recovery from AUD (Seo et al., 2013). We observed the opposite effect in our cohort, where increased rs-FC between SN and DMN was more likely to be predictive of the low-sip group. However, the low-sip group started at a higher level of alcohol sipping compared to the high-sip group. Thus, hyperconnectivity at baseline between the SN and DMN might be indicative of this gap rather than the trajectory of sipping over time. Additionally, another adult study exploring alterations in rs-FC of alcohol-users identified that alcohol-users exhibited decreased functional connectivity in regions related to the sensory and motor network (Vergara et al., 2017). Similarly, hypoconnectivity between SMN and right amygdala or right cerebellum was representative of the low-sip group, while hyperconnectivity between SMN and right nucleus accumbens was representative of the high-sip group. While the individuals in both groups reported alcohol sipping at the beginning of the study, these findings might differentiate between those who continue sipping and those who decide to reduce their sipping over time. Although the age and the level of alcohol consumption in our cohort is not comparable to alcohol-dependent patients, these findings can underline potential differences between alcohol dependence and alcohol experimentation.

There is a growing need to analyze functional connectivity interactions in different alcohol and substance use patterns as children transition into adolescence (Huntley et al., 2020; Kardan et al., 2024). Similarly to the development of brain structure, brain function evolves throughout life. During neurodevelopment, functional networks become organized through segregation and integration, while each functional network maturates at a different pace (Edde et al., 2021). During early adolescence, brain regions responsible for basic functions start to strengthen, while regions involved in complex tasks undergo dynamic changes in connectivity, with connections either strengthening or weakening between the ages of 14 and 26 (Váša et al., 2020). In our analysis, we found stable interactions between rs-FC between SMN, RTN and FPN and subcortical regions related to memory, learning, motivation, and sensory and motor outputs in the differentiation of the sipping trajectories of alcohol initiators. These findings suggest that early functional connectivity interactions may contribute to alcohol use patterns during adolescence. Nevertheless, this area is understudied and needs further replication in children aged 7-14.

The main takeaway from this study is that different factors could impact distinct trajectories of alcohol sipping patterns in children. The contrast between individuals who abstained from alcohol initiation and those who exhibited an increasing trajectory of alcohol sipping over time was primarily explained by attitudes toward drinking, family and school environment, brain structure, and functional connectivity. Additionally, among those who experimented with alcohol at the start of the study, the distinction between those who continued sipping and those who reduced their sipping over time was influenced by family environment, genetic ancestry, and functional connectivity, but not by brain structure. These findings highlight the importance of early interventions and preventions to reduce the risk of problematic drinking behaviors. At the individual level, adopting healthy attitudes towards alcohol exposure and improving self-regulation skills may help prevent early initiation. At the family and community levels, promoting supportive environments, setting clear expectations around alcohol use, and implementing educational programs on the risks of early alcohol exposure could be effective. Additionally, understanding the neural and genetic influences on drinking trajectories can inform targeted prevention strategies for at-risk youth, contributing to healthier developmental outcomes.

This study has several limitations. First, although the best models outperformed random guessing, classifying individuals who initiated alcohol use but continued drinking versus those who chose to abstain remains challenging due to population heterogeneity. Second, the data-driven approach we used carries a risk of overfitting, especially when dealing with complex data. Additionally, we relied on complete data and on baseline variables available in the ABCD study and did not use any imputation techniques to address missing variables, which limited our sample size. Third, although the study aimed to use a data-driven approach, the input data included in Aim 1 was limited by the multilevel, multidomain factors previously identified in studies on alcohol initiation and binge drinking.

In conclusion, we applied machine learning models to identify multilevel, multidomain, multimodal neuroimaging features predictive of alcohol sipping patterns as children transition into adolescents. These factors not only predicted alcohol sipping patterns, but also suggested that multilevel, multidomain factors can interact with brain structures and functions prior to alcohol initiation. Nevertheless, further replication studies on other young adolescent cohorts need to be conducted to validate the current findings. Given that few studies match the size and comprehensiveness of the ABCD study, our selected features can be categorized similarly to our aims, facilitating comparisons with smaller datasets. Future research using the ABCD dataset could focus on identifying new alcohol sipping trajectories and drinking patterns as adolescents age, increase their alcohol exposure, and as additional time points become available. Moreover, assessing individual differences within subgroups representative of alcohol use behaviors over time could provide deeper insights into the underlying risk and protective factors, helping to refine prevention and intervention strategies tailored to specific subgroups.

## Data Availability

Data used in the preparation of this article were obtained from the Adolescent Brain Cognitive DevelopmentSM (ABCD) Study (https://abcdstudy.org), held in the NIMH Data Archive (NDA). This is a multisite, longitudinal study designed to recruit more than 10,000 children age 9-10 and follow them over 10 years into early adulthood. The ABCD Study is supported by the National Institutes of Health and additional federal partners under award numbers U01DA041048, U01DA050989, U01DA051016, U01DA041022, U01DA051018, U01DA051037, U01DA050987, U01DA041174, U01DA041106, U01DA041117, U01DA041028, U01DA041134, U01DA050988, U01DA051039, U01DA041156, U01DA041025, U01DA041120, U01DA051038, U01DA041148, U01DA041093, U01DA041089, U24DA041123, U24DA041147. A full list of supporters is available at https://abcdstudy.org/federal-partners.html. A listing of participating sites and a complete listing of the study investigators can be found at https://abcdstudy.org/consortium_members/. ABCD consortium investigators designed and implemented the study and/or provided data but did not necessarily participate in the analysis or writing of this report. This manuscript reflects the views of the authors and may not reflect the opinions or views of the NIH or ABCD consortium investigators. The ABCD data repository grows and changes over time. The ABCD data used in this report came from DOI: 10.15154/z563-zd24. Additional support for this work was made possible from NIEHS R01-ES032295 and R01-ES031074.

