## Supplemental Materials for "Integrating Multilevel, Multidomain and Multimodal Neuroimaging Factors to Predict Early Alcohol Exposure Trajectories Using Explainable AI"

Supplemental Material

1. Measures
2. Table S1: Model performances for the no-vs-high-sip contrast with multilevel, multidomain features as input data.
3. Table S2. Model performances for the low-vs-high-sip contrast with multilevel, multidomain features as input data
4. Table S3. Model performances for the no-vs-high-sip contrast with structural features and functional connectivity as input data.
5. Table S4. Model performances for the low-vs-high-sip contrast with functional connectivity as input data.
6. Fig S1. Accumulated local effects (ALE) plots of main effects in the classification of low-sip vs. high-sip based on the Elastic Net model.
7. Fig S2. Accumulated local effects (ALE) plots of second-order interaction effects in the classification of no-sip vs. high-sip, after accounting for main effects.
8. Fig S3. Accumulated local effects (ALE) plots of second-order interaction effects in the classification of low-sip vs. high-sip, after accounting for main effects.
1. Measures

*Individual level:*

*Family history of drug use* was assessed at baseline as if any of the participant’s blood relative has ever been to the doctor or a counselor about any emotional or mental problems, or problems with alcohol or drugs. *Prenatal drug exposure* was recorded for each of the substances the mother was exposed to during pregnancy. *BMI* was calculated as the ratio of weight (in kg) and height (in meters) squared, where values less than 10 and greater than 40 were considered outliers and were excluded from the analysis (Center for Disease Control and Prevention, 2022). *Genetic ancestry* was captured using 32 principal components of genetic ancestry derived from genotype data. *Sleep time* was measured by the parent as the child’s total number of hours of sleep per night. *Physical activity* was measured based on the Sports and Activities Involvement Questionnaire (SAI-Q) including information on 23 sports. The physical activity variable was computed as the mean participation per sport as hours per week (Palmer et al., 2021). The sum of the physical activity for each sport represented the total average time spent participating in sports. *Weekend and weekday screen time* was measured as the number of hours spent using a screen on activities not related to schoolwork, such as watching TV, texting, using social media, playing video games etc.

*Internalizing, externalizing and conduct problems* were assessed using the Child Behavior Checklist (CBCL) scale, as reported by the parent. Internalizing symptoms include depression, anxiety, and withdrawal, while externalizing symptoms encompass hyperactivity, aggression, and non-compliance. Conduct problems are a subset of externalizing problems that are more severe and persistent. Higher scores indicated more parent-reported problems. *Childhood trauma* was measured through the parent-reported Kiddie Schedule for Affective Disorders and Schizophrenia (KSADS) and was represented through 4 variables: the total number of lifetime traumatic events, the total number of post-traumatic stress disorder (PTSD) symptoms, past or present, PTSD diagnosis, past or present, based on the Diagnostic and Statistical Manual of Mental Disorders Fifth Edition (DSM-5) and PTSD that did not fulfill the DSM-5 criteria, past or present (American Psychiatric Association, 2013).

*Personality traits* were assessed using the UPPS-P for Children Short Form, which measures impulsivity through subscales for negative urgency, positive urgency, sensation seeking, lack of planning, and lack of perseverance. Additionally, the behavioral inhibitory/approach systems (BIS/BAS) scales were used to assess BIS and BAS subscales for reward responsiveness, drive, and fun seeking. Higher scores indicated a higher propensity for the specific trait. The child's *prosocial behavior* was assessed as the mean of three items from the self-reported Strengths and Difficulties Questionnaire (SDQ), with higher scores indicating higher levels of prosocial behavior (Goodman et al., 1998).

Participants also reported information if they have ever had a *puff of tobacco or marijuana* and a *sip of alcohol* before the start of the study. For participants who reported alcohol sipping at baseline, the *age of the first alcohol* sip in years was also included in the analysis. *Positive alcohol expectancies (PAE)* were measured based on the Alcohol Expectancies Questionnaire – Adolescent, Brief (AEQ-AB). Since this variable was unavailable at baseline, we used the data from the 1-year follow-up, as the values remained constant across the following three time points. *Alcohol curiosity and intent* were measured based on what extent the participant has ever been curious to try alcohol, and on the intention to try alcohol soon, respectively.

*Family level:*

*Family income* was measured as the total combined family income before taxes in the past year. The *highest parental education* level was measured as the highest education level of the household. *Parental behavior*, also known as emotional neglect, was reported by the child and measured as the total sum of the scores from both caregivers. In case the household was formed by only one caregiver, then parental behavior was measured based on the only caregiver’s total score. Higher total score reflected more positive behavior towards the child. *Parental monitoring* was assessed using the mean parental monitoring score (PMS), and a higher PMS suggested greater levels of parental monitoring and involvement. *Family conflict* was assessed using the Family Environment Scale (FES) as the total scores reported by the parent regarding family relationships at home, where higher scores indicated a greater level of conflict within the family. *Parental rules* regarding the child’s alcohol consumption were reported by the parent. Parents were also asked if they have *penalties* for not respecting the rules regarding youth alcohol consumption in the household. *Alcohol availability* to the child was also reported.

*Community level:*

*Neighborhood disadvantage* was measured by the Area Deprivation Index (ADI). Higher ADI indicates a higher level of disadvantage. *Neighborhood safety* was measured assessing the feeling of being safe in the neighborhood. Higher neighborhood safety scores reflected a safer environment. *Area type* categorized as urban, urban cluster or rural was also reported based on the Census Tract Urban Classification on the primary address. Parents also reported the child’s *religion* at baseline. *Academic performance* was measured by the average grades reported by parents. *School environment, involvement and disengagement* were measured using the School Risk and Protective Factors questionnaire (SRPF). Higher scores reflected a better school environment, more involvement in school and more disengagement, respectively. Participants were also asked to report the *number of friends drinking alcohol* at baseline.

**Table S1. Model performances for the no-vs-high-sip contrast with multilevel, multidomain features as input data.**

| **Model** | **AUC**  **(95% CI)** | **Sensitivity** | **Specificity** |
| --- | --- | --- | --- |
| CART | 0.673  (0.639, 0.707) | 0.520 | 0.852 |
| Random Forest | 0.683  (0.649, 0.717) | 0.547 | 0.818 |
| **Ridge** | **0.707**  **(0.674, 0.740)** | **0.646** | **0.768** |
| Elastic Net | 0.697  (0.664, 0.731) | 0.623 | 0.771 |
| LASSO | 0.6883  (0.6546,0.7221) | 0.596 | 0.780 |
| SVM linear kernel | 0.670  (0.636, 0.704) | 0.525 | 0.816 |
| SVM 2^nd^ order polynomial kernel | 0.680  (0.646, 0.714) | 0.557 | 0.804 |
| SVM 3^rd^ order polynomial kernel | 0.678  (0.644,0.712) | 0.520 | 0.835 |
| SVM 4^th^ order polynomial kernel | 0.632  (0.599,0.665) | 0.386 | 0.878 |
| SVM 5^th^ order polynomial kernel | 0.588  (0.559,0.617) | 0.247 | 0.930 |
| SVM radial kernel | 0.683  (0.649, 0.717) | 0.565 | 0.802 |
| Ensemble learning | 0.698  (0.664,0.731) | 0.614 | 0.781 |

**Table S2. Model performances for the low-vs-high-sip contrast with multilevel, multidomain features as input data**

| **Model** | **AUC**  **(95% CI)** | **Sensitivity** | **Specificity** |
| --- | --- | --- | --- |
| CART | 0.508  (0447, 0.569) | 0.473 | 0.543 |
| Random Forest | 0.559  (0.499, 0.620) | 0.512 | 0.606 |
| Ridge | 0.575  (0.514, 0.636) | 0.586 | 0.564 |
| Elastic Net | 0.581  (0.520, 0.641) | 0.640 | 0.521 |
| LASSO | 0.587  (0.526, 0.647) | 0.631 | 0.543 |
| SVM linear kernel | 0.523  (0.461, 0.584) | 0.505 | 0.543 |
| SVM 2^nd^ order polynomial kernel | 0.543  (0.482, 0.604) | 0.596 | 0.489 |
| SVM 3^rd^ order polynomial kernel | 0.505  (0.495, 0.516) | 1 | 0.011 |
| SVM 4^th^ order polynomial kernel | 0.500  (0.488, 0.513) | 0.990 | 0.011 |
| SVM 5^th^ order polynomial kernel | 0.503  (0.491, 0.514) | 0.995 | 0.011 |
| SVM radial kernel | 0.549  (0.488, 0.610) | 0.512 | 0.585 |
| **Ensemble learning** | **0.590**  **(0.536, 0.645)** | **0.404** | **0.777** |

**Table S3.** **Model performances for the no-vs-high-sip contrast with structural features and functional connectivity as input data.**

| **Model** | **AUC**  **(95% CI)** | **Sensitivity** | **Specificity** |
| --- | --- | --- | --- |
| CART | 0.587  (0.556, 0.618) | 0.679 | 0.495 |
| Random Forest | 0.572  (0.539, 0.604) | 0.484 | 0.659 |
| Ridge | 0.570  (0.537,0.602) | 0.512 | 0.627 |
| **Elastic Net** | **0.621**  **(0.589, 0.652)** | **0.651** | **0.591** |
| LASSO | 0.583  (0.551, 0.616) | 0.538 | 0.639 |
| SVM linear kernel | 0.584  (0.552, 0.617) | 0.559 | 0.609 |
| SVM 2^nd^ order polynomial kernel | 0.515  (0.494, 0.536) | 0.889 | 0.141 |
| SVM 3^rd^ order polynomial kernel | 0.501  (0.497, 0.505) | 0.004 | 0.997 |
| SVM 4^th^ order polynomial kernel | 0.499  (0.498, 0.500) | 0.000 | 0.999 |
| SVM 5^th^ order polynomial kernel | 0.500  (0.494, 0.506) | 0.008 | 0.992 |
| SVM radial kernel | 0.501  (0.492, 0.511) | 0.024 | 0.979 |
| Ensemble Learning | 0.566  (0.534, 0.598) | 0.587 | 0.545 |

**Table S4. Model performances for the low-vs-high-sip contrast with functional connectivity as input data.**

| **Model** | **AUC**  **(95% CI)** | **Sensitivity** | **Specificity** |
| --- | --- | --- | --- |
| CART | 0.493  (0.439, 0.547) | 0.526 | 0.460 |
| Random Forest | 0.452  (0.402, 0.502) | 0.757 | 0.339 |
| Ridge | 0.525  (0.471, 0.579) | 0.518 | 0.532 |
| **Elastic Net** | **0.560**  **(0.506, 0.613)** | **0.530** | **0.589** |
| LASSO | 0.549  (0.496, 0.613) | 0.518 | 0.581 |
| SVM linear kernel | 0.505  (0.451, 0.559) | 0.470 | 0.540 |
| SVM 2^nd^ order polynomial kernel | 0.500  (0.499,0.510) | 0.992 | 0.008 |
| SVM 3^rd^ order polynomial kernel | 0.534  (0.496,0.572) | 0.899 | 0.169 |
| SVM 4^th^ order polynomial kernel | 0.496  (0.481, 0.511) | 0.976 | 0.016 |
| SVM 5^th^ order polynomial kernel | 0.500  (0.500, 0.500) | 1.000 | 0.000 |
| SVM radial kernel | 0.550  (0.499,0.601) | 0.729 | 0.371 |
| Ensemble Learning | 0.495  (0.442, 0.5481) | 0.397 | 0.613 |

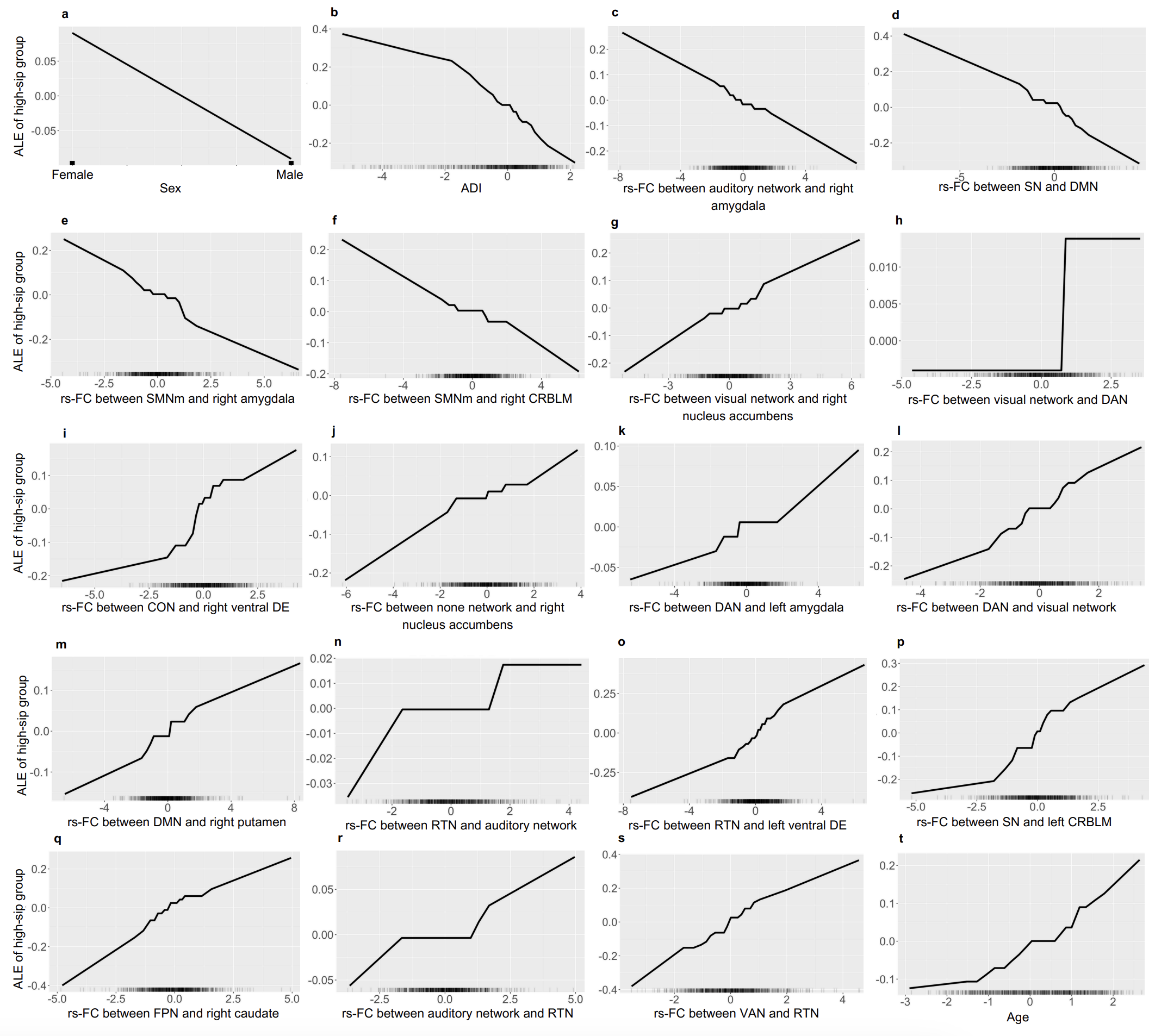

**Fig S1. Accumulated local effects (ALE) plots of main effects in the classification of low-sip vs. high-sip based on the Elastic net model.**

*Note: ADI – area deprivation index, SN – salience network, DMN – default mode network, SMNm – sensorimotor mouth network, CRBLM – cerebellum cortex, DAN – dorsal attention network, CON – cingulo-opercular network, DE – diencephalon, RTN – retrosplenial temporal network, FPN – frontoparietal network, VAN – ventral attention network*

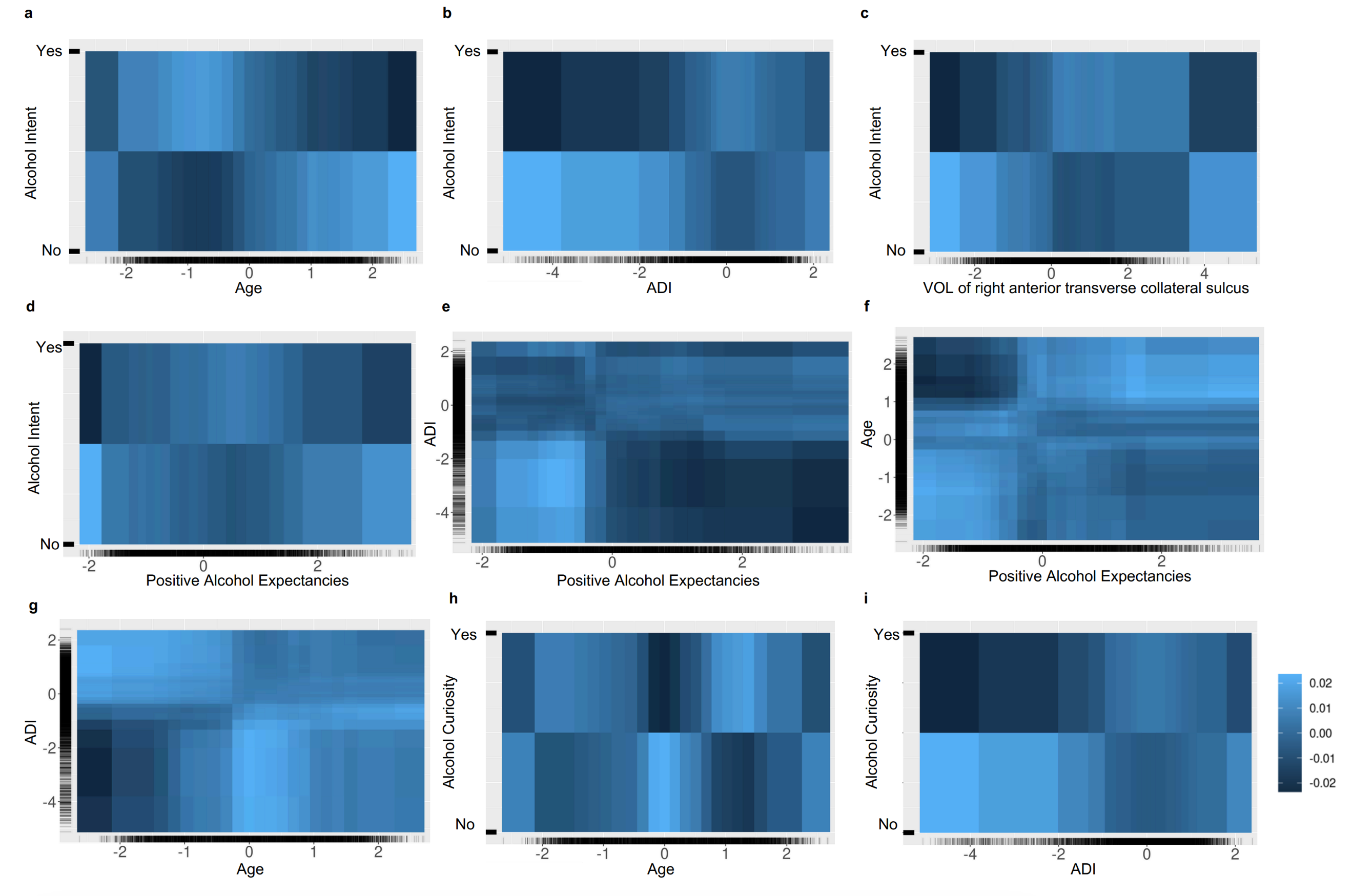

**Fig S2. Accumulated local effects (ALE) plots of second-order interaction effects in the classification of no-sip vs. high-sip, after accounting for main effects.**

*Note: Lighter shade represents an above average effect, and darker shade represents a below average prediction on the likelihood of belonging to the high-sip group, accounting for all other main effects.*

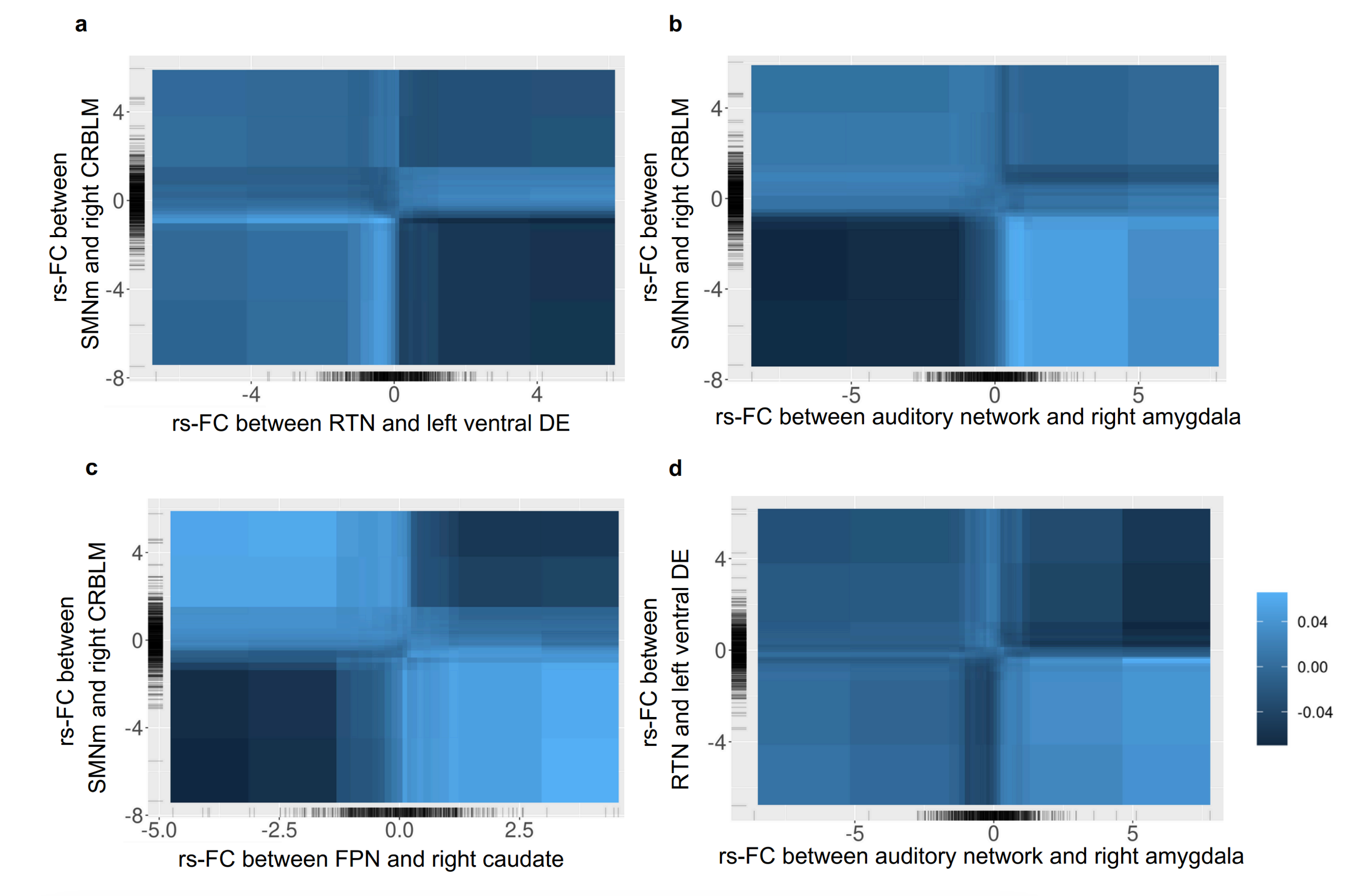

**Fig S3. Accumulated local effects (ALE) plots of second-order interaction effects in the classification of low-sip vs. high-sip, after accounting for main effects.**

*Note: Lighter shade represents an above average effect, and darker shade represents a below average prediction on the likelihood of belonging to the high-sip group, accounting for all other main effects.*
